# Real time scalable data acquisition of COVID-19 in six continents through PySpark - a big data tool

**DOI:** 10.1101/2021.07.04.21259983

**Authors:** Tanvi S. Patel, Daxesh P. Patel, Chirag N. Patel

## Abstract

Severe acute respiratory syndrome coronavirus 2 (SARS-CoV-2) was declared as a global emergency in January 2020 due to its pandemic outbreak. To examine this Coronavirus disease 2019 (COVID-19) effects various data are being generated through different platforms. This study was focused on the clinical data of COVID-19 which relied on python programming. Here, we proposed a machine learning approach to provide a insights into the COVID-19 information. PySpark is a machine learning approach which also known as Apache spark an accurate tool for the searching of results with minimum time intervals as compare to Hadoop and other tools. World Health Organization (WHO) started gathering corona patients’ data from last week of the February 2020. On March 11, 2020, the WHO declared COVID-19 a global pandemic. The cases became more evident and common after mid-March. This paper used the live owid (our world in data) dataset and will analyse and find out the following details on the live COVID-19 dataset. (1) The daily Corona virus scenario on various continents using PySpark in microseconds of Processor time. (2) After the various antibodies have been implemented, how they impact new cases on a regular basis utilizing various graphs. (3) Tabular representation of COVID-19 new cases in all the continents.

## Introduction

In early December 2019, an unidentified virus known as coronavirus was discovered in Wuhan, China, among a group of patients suffering from severe tuberculosis, and it quickly spread to other cities. The modern infection severe intense respiratory syndrome coronavirus 2 (SARS-CoV-2) was declared by the World Health Organization, and the illness caused by this unused infection was called coronavirus disease 2019 (COVID-19)(You et al. 2020)(Patel et al. 2020).

In January 2020, the World Health Organization (WHO) declared a global emergency, and the situation quickly escalated, prompting WHO to declare COVID-19 a global pandemic on March 11, 2020 (Cucinotta and Vanelli 2020; Patel et al. 2021a). COVID-19 is spread predominantly from person to person, most commonly through respiratory droplets from coughing, sneezing, or talking. All the researchers, biologist, Pharmaceuticals and doctors were trying to find the medicine, vaccines and treatment to control COVID-19 (Patel et al. 2021b). In December 2020, The U.S. Food and Drug Administration (USFDA) granted an Emergency Use Authorization (EUA) for the Pfizer-BioNTech COVID-19 Vaccine. The FDA granted an EUA for the use of the Moderna COVID-19 Vaccine on December 18, 2020. The present study provides an insights of the COVID-19 new cases curve after antibody vaccines were introduced using machine learning approach (Ong et al. 2020).

Since MS Excel has a 255-row limit for processing and visualizing data, we can use tableau, Hadoop MapReduce, or Apache Spark for big data analysis (Gupta et al. 2017). However, when compared to other machine learning tools, Apache Spark provides us with quick, reliable, and accurate results. Apache spark can also handle Parquet, ORC, JSON, CSV, and text format. There are many ways of data visualization and analysis, such as theoretical, graphical and tabular forms. Graphs are striking psychological resources because they rapidly and efficiently display information. Often, when provided by a graph, data can be better interpreted than by a hypothesis since a pattern or comparison can be unveiled by the graph and chart. The Python scripting language offers numerous libraries, such as pandas, to construct very powerful graphs and charts (Gupta et al. 2017; Stancin and Jovic 2019). PySpark includes comprehensive and quality ways to run zeta bytes and peta bytes of data on parallel clusters a lot faster than conventional python applications for machine learning applications. PySpark offers methods for statistical analysis and a ML (Machine Learning) pipeline. In specific, Apache spark is emerged as popular open framework that support the various data visualization techniques like different charts (Bar, line, pie, etc). Apache Spark provides JAVA, Scala, R and python API’s but among all of these, python is simple scripting language and good for machine learning due to its versatile existence. Python’s popularity in the field of data science has reached new heights, especially in terms of freely accessible frameworks and libraries. Among the many free libraries available, pandas, NumPy, and Matplotlib are well-known and widely used in the field of data analysis (Ghaffar et al. 2015; Assefi et al. 2017; Gupta et al. 2017; Magar et al. 2021).

In this COVID-19 challenge, everyday millions of data is being generated and these data are continuously employed to established different preventions and control steps. COVID-19 datasets are increasing in size day by day as well as it is becoming sophisticated too. It is necessary to build solutions using statistical methods to leverage this wealth of data. Due to the rise in data and new data on a regular basis, new scripting codes every day need to be analysed to create new visualizations of the data as well as new graphs. In order to solve this issue, this paper incorporates live data analysis so that visualization is often updated every day as data changes and there is no need for new scripts every day. This study focuses on the daily cases and daily deaths in various continents due to COVID-19 and makes statistical graphs for all continents. This work also highlights confirmed cases and death ratios in a tabular format, which allows for easy comparison of the entire world’s continents in a single table. Using PySpark, compare all the Earth’s continents in a graph and table.

## Methodology

### Data curation

In order to perform the statistical analysis, the data of COVID-19 collected from all over in world. It includes confirmed cases, new cases, deaths, hospitalizations, testing, and vaccinations, throughout the duration of the COVID-19 pandemic. COVID-19 cases and death were collected from COVID-19 Data Repository by the Center for Systems Science and Engineering (CSSE) at Johns Hopkins University (Badr et al. 2020; Magar et al. 2021). European countries vaccination against COVID-19 details were obtained from the European Surveillance System (TESSy) webpage (Ammon and Faensen 2009). Vaccinations against COVID-19 data was retrieved by the Data team from official reports of all across the globe. Other data was selected from a variety of sources like United Nations, World Bank, Global Burden of Disease, Blavatnik School of Government and more. Web-based application was used for the utilization of the COVID-19 data with live monitoring (Ghaffar et al. 2015). Figure-1 depicts the workflow of proposed study which comprises of various data collection and administered. The raw COVID-19 data was customized using text editor with normalization with the filtered by continent accountability.

**Figure-1.**
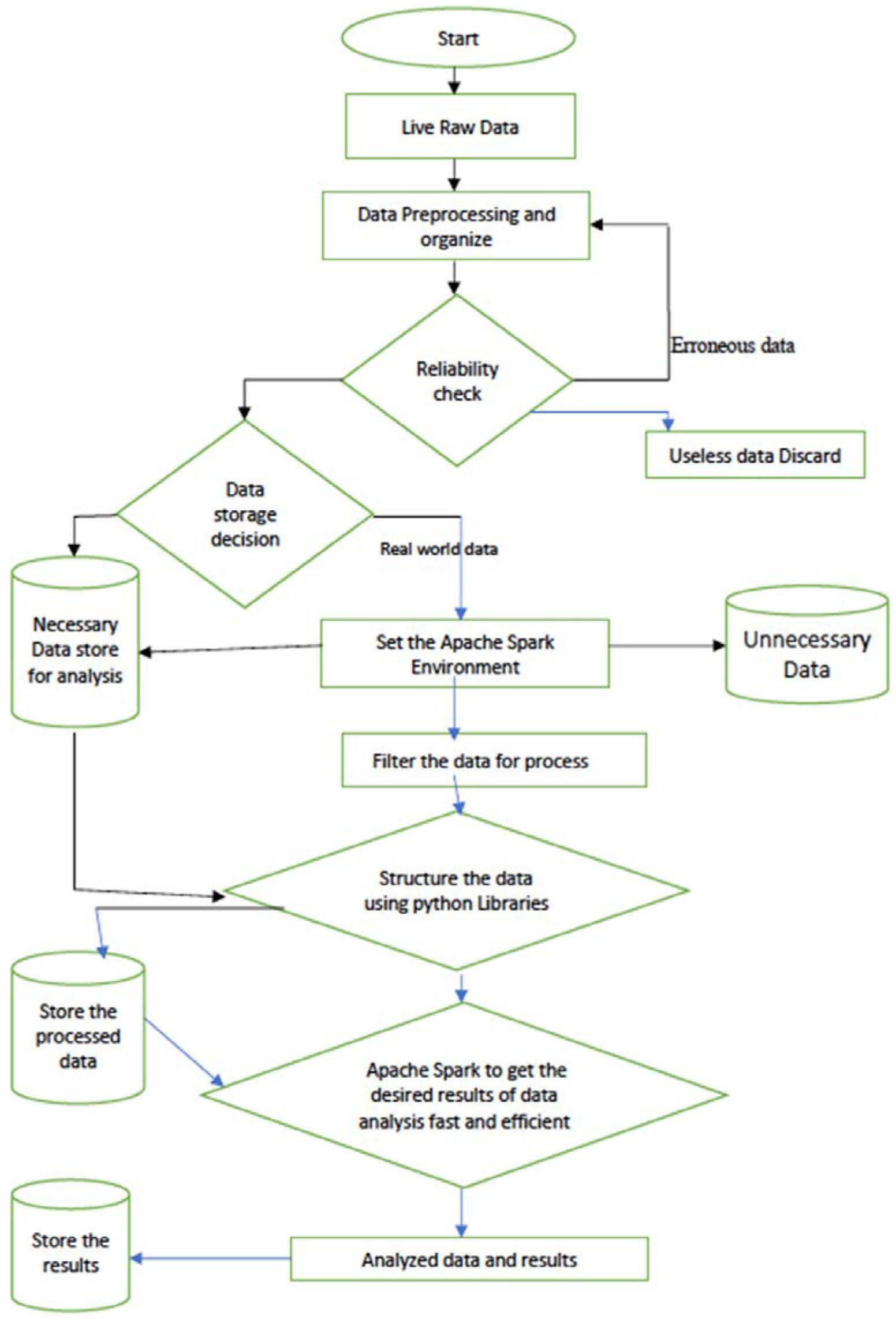
Flowchart of proposed study.

### Machine learning Approach: Apache Spark with API python

Apache Spark is a fast and trustable general-purpose cluster computing framework. This framework offers API (application programming interfaces) in Java, Python, Scala and R script. Spark is an Apache incubator cluster computing system that is focused in making data processing easier with top agile application in coding as well as running. It is very fast at both running and coding (Salloum et al. 2016; Assefi et al. 2017). This applicability was applied for the current study which proposed novel data structure of COVID-19 using Spark core and upper-level libraries such as Spark’s MLlib for machine learning, GraphX for graph analysis, Spark Streaming for stream processing, and Spark SQL for structure data processing make up Apache Spark (Assefi et al. 2017; Gupta et al. 2017). Real-time data, such as streaming, video, and social media data, was also analysed by Apache Spark. The promising and accurate data processing was performed through Apache Spark which includes the 7500 rows at a single instance or operation with meticulously. Figure-2 shows the Apache spark architecture incorporates four APIs with correlation to COVID-19 dataset to analyse it on case by case basis with numbers of deaths and vaccines are being employed till now. The average speed of the Apache Spark is 75 thousand rows and 59 columns in 4.46 μs of total time and 589 μs of wall time (Binti Hamzah et al. 2020).

**Figure-2.**
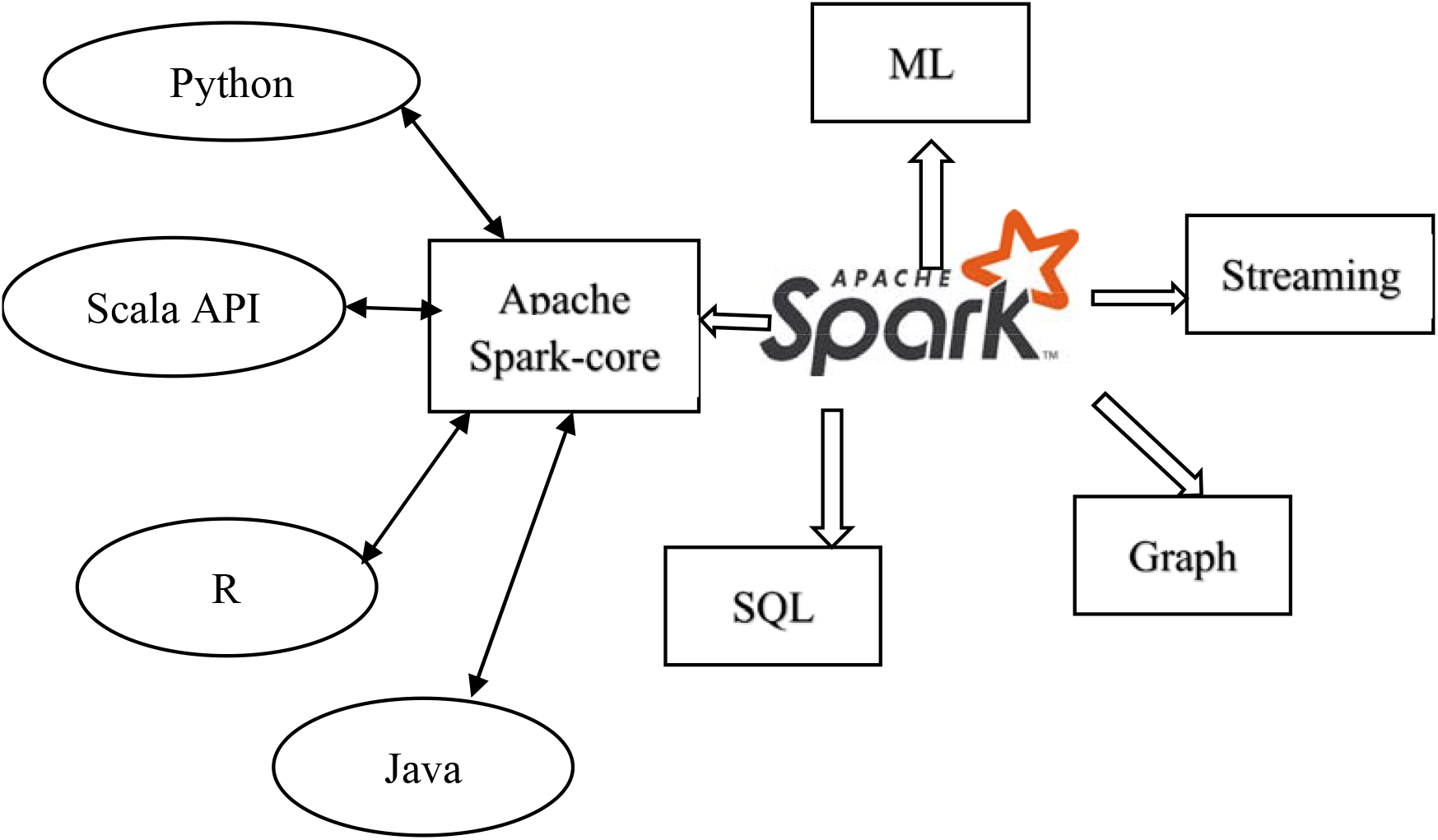
Apache Spark and API (Application Programming Interface).

Python’s prominence is increasing, widely in the all areas like Bio informatics and data analysis as well as in programming too (Latif et al. 2020). As a result, the number of free frameworks and libraries for use is growing. Various Python big data research libraries, such as pandas, matplotlib, NumPy, and seaborn, are also used in this analysis (Figure-3). Python libraries are commonly used to visualize data, and visualization tool is becoming simpler with the use of these free libraries.

**Figure-3.**
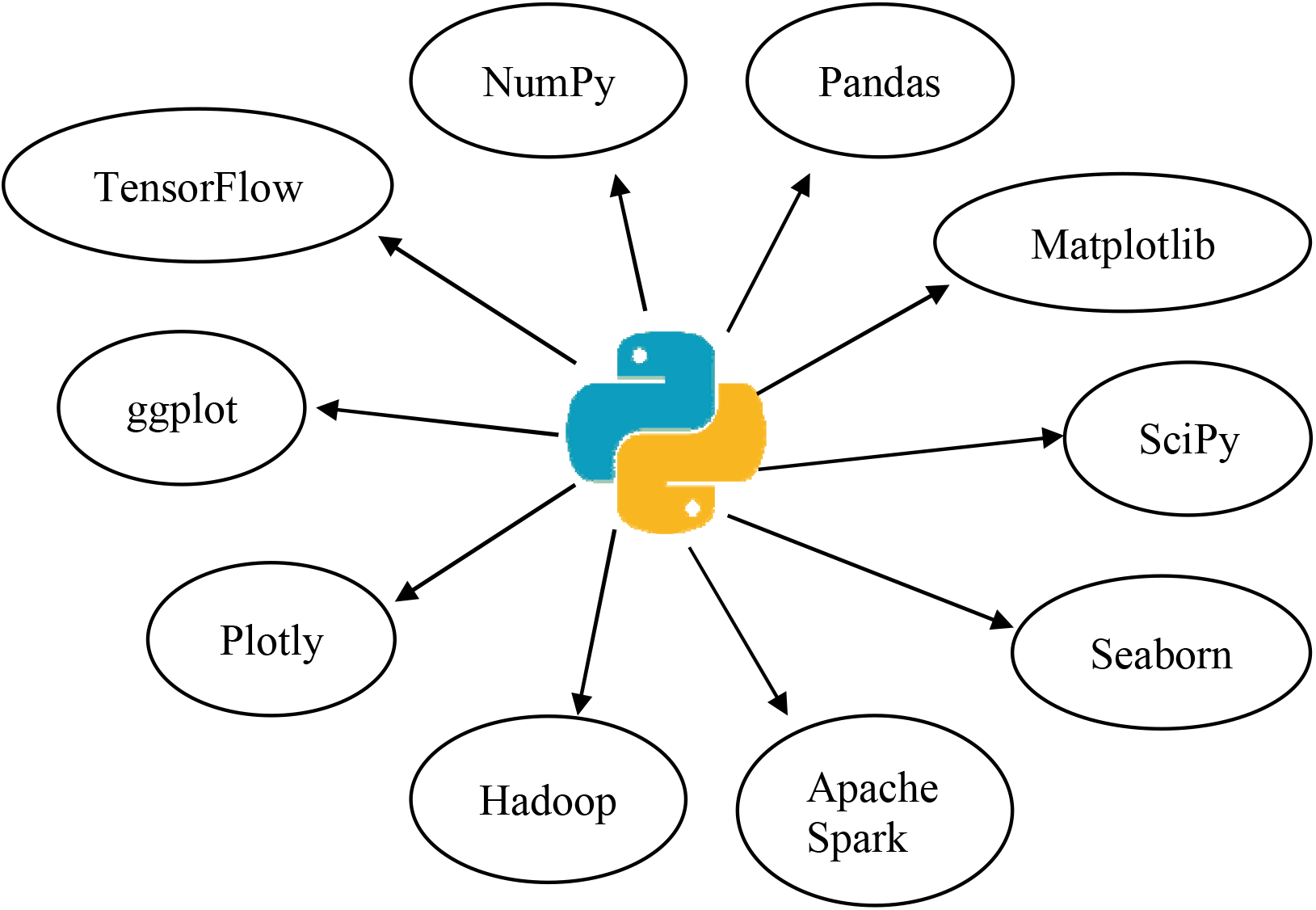
Python and it’s used libraries.

***Formulas***

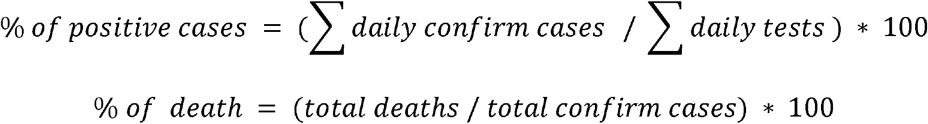

### Script used to find charts

The given script shows how a small amount of code can create a graph with real time data rows of data in a short amount of time. Real time data set was obtained from https://raw.githubusercontent.com/owid/covid-19-data/master/public/data/owid-covid-data

.csv. PySpark Scripting code was uploaded on https://github.com/pateltanvi2992/Data-Analysis-of-Covid-19-using-Spark-(Patel2021).

## Results and Discussion

The vast volume of daily produced data has pushed us beyond the limits of conventional processing capacities, necessitating the adoption of advanced computing infrastructures that can process data in parallel and distributed ways. The generated data consists of huge in size and requires innumerable memory space, so Apache Spark was adopted to rectify this problem. The Apache spark platform and its library provides the exploration of qualitative and quantitative attributes collection. With the help of Python large-scale real-world data experiments of COVID-19 were applied for this study. These data were represented in six different continents.

Given demographic table-1 is describe the whole scenario of the COVID-19 in six continents. The limitation of this study in our analysis we did not consider data from the seventh continent Antarctica as there is not any permeant resident staying although there is at least 36 people confirmed with COVID infection (https://www.barrons.com/; https://www.coolantarctica.com/).

**Table-1.** Covid-19 Tabular data (till May-15^th^-2021).

| Continents | Population | Total Tests | Positive cases | Total recover | People Vaccinated |
| --- | --- | --- | --- | --- | --- |
| Asia | 4,669,823,825<br>(60.8%) | 508,287,012 | 42,734,316 | 42,157,308 | 624,708,871 |
| North America | 368,869,647<br>(4.8%) | 484,569,026 | 38,505,833 | 37,637,694 | 310,026,856 |
| South America | 433,340,178<br>(5.6%) | 46,209,536 | 27,164,684 | 26,430,452 | 89,405,489 |
| Europe | 746,400,000<br>(9.7%) | 662,528,505 | 46,740,089 | 45,688,120 | 255,813,054 |
| Oceania | 43,066,356<br>(0.56%) | 12,149,407 | 46,922 | 45,851 | 3,501,157 |
| Africa | 1,364,509,774<br>(17.78%) | 29,036,507 | 4,716,496 | 4,582,390 | 13,949,677 |

### Asia

Asia has a total of 23225625.0 COVID-19 positive cases and 22836076.0 recovered cases as of May 15th, 2021. According to this, the mortality rate will be 1.6359 percent until May 15th, 2021. From February 24, 2020 to May 15th, 2021, the daily recover cases and daily confirm cases are depicted in Figure-4 (B) (scatter graph). On September 16, 2020, the highest number of new cases were registered in India, with 97894.0 (new cases) out of 1136613.0 (new tests). The COVID-19 cases peaked in September 2020, according to Figure-5 (B) (histogram), and then started to decline. Total tests performed in Asia were 373625749.0, with 23225625.0 confirmed cases. As a result of this, 6.2163 percent of the cases tested positive.

**Figure-4.**
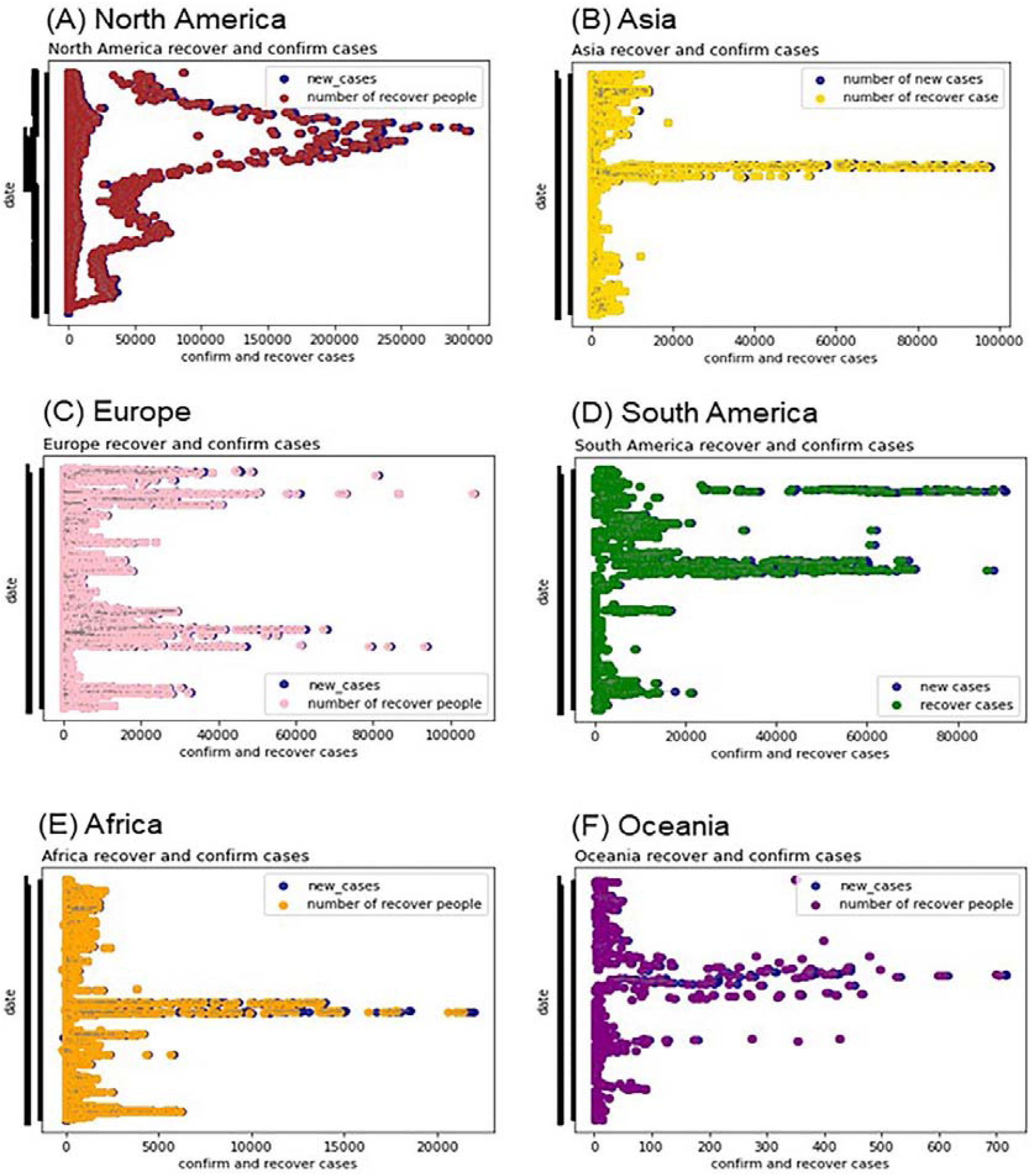
COVID-19 cases distribution based on recovery rate from various continents.

### Africa

Africa has total COVID-19 positive cases till May 15th 2021 are 4027046.0 and recover are 3912657.0. On basis of that the death ratio is 2.6701 %. Figure-4 (E) is showing the daily recover cases and positive cases. Figure-5 (E) is showing the positive cases going from 25th February 2020 to May 15th 2021. Although Africa has 17.2% of the world’s population, it only has about 5% of COVID-19 cases diagnosed and 3% of COVID-19-related mortality (Bamgboye et al. 2020).

**Figure-5.**
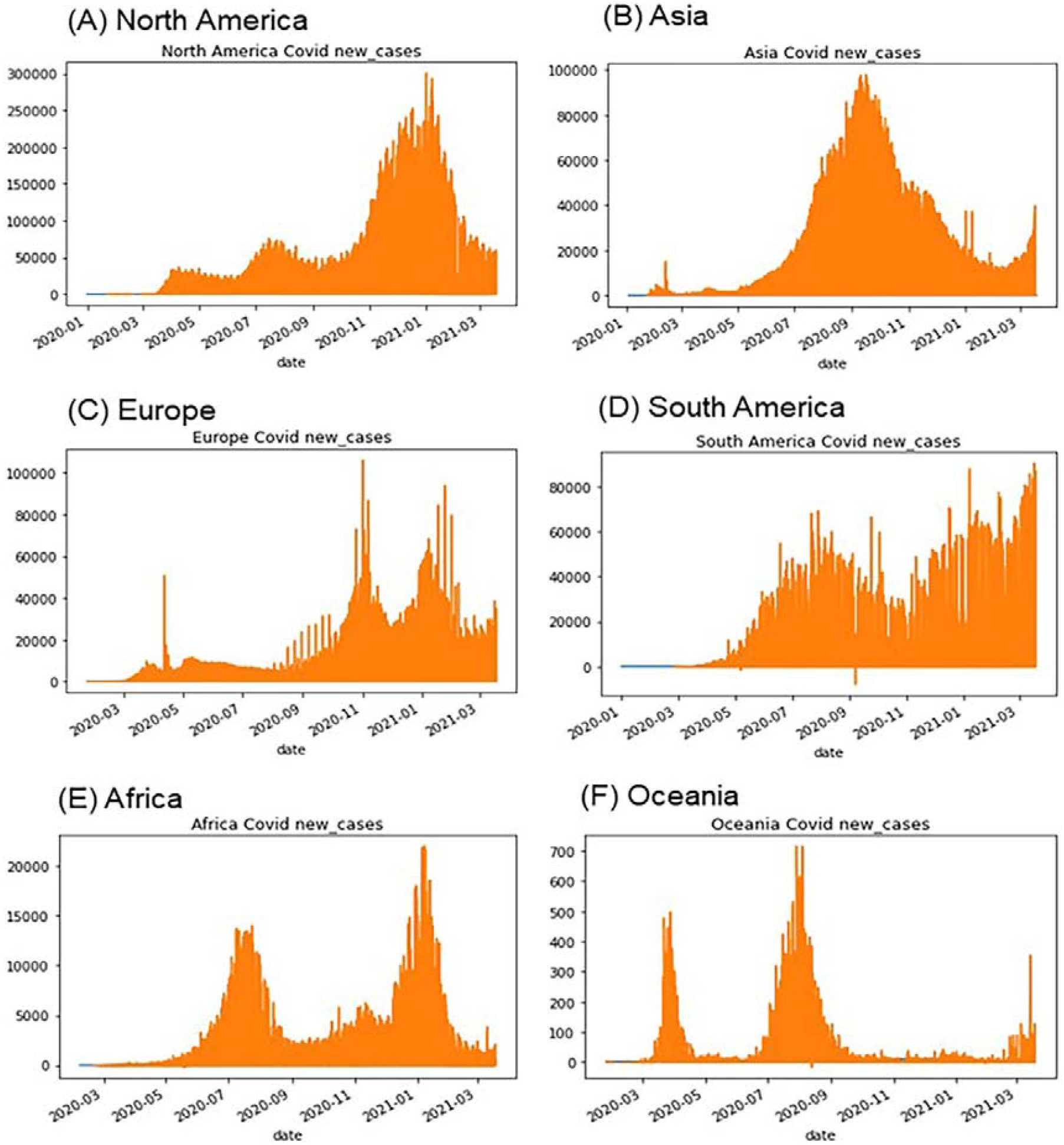
Histographic representation of COVID-19 updated cases in various regions.

### Europe

Europe has total COVID-19 positive cases till May 15th 2021 are 36470789.0 and recover cases are 35620243.0. On basis of that the death ration in Europe was 2.3321%. The maximum cases reported on 2nd November 2020 in France which was 106091.0. Figure**-4** (C) shows the recover cases and positive cases through the scatter graph. The total test occur in Europe was 448349376.0. Figure-5 (C) is showing the graph of positive cases from 11th March 2020 to 13th March 2021.

### North America

North America has total COVID-19 positive cases till May 15th, 2021 are 33850076.0 and recover cases are 33070581.0. The percentage of the death ratio in North America was 2.29889 %. Figure-4 (A) (scattered graph) is showing the daily new cases and daily recover cases from 24th February 2020 to May 15^th^, 2021. The maximum new confirm cases reported in North America on 2nd January 2021 in United states was 300416.0 among 108349.0 new tests. The total test occurred in North America till May 15^th^, 2021 was 363400300.0. So, 9.3148% positive cases among all tests. Figure-5 (A) (histogram) is showing the cases going from February 24th, 2020 to May 15^th^, 2021 (Control 2021).

### South America

South America has total COVID-19 positive cases till May 15^th^, 2021 are 19191051.0 and recover cases are 18693169.0. Based on this the death ratio of the South America was 2.5853%. Figure-4 (D) (scatter graph) is showing the daily new cases and daily recover cases. The maximum new cases reported in South America 87843.0 in Brazil on 7^th^, January 2021. The total test occur in South America was 4245255498.0. Figure-5 (D) is showing the graph going for daily positive cases in South America from 1st January 2020 to May 15^th^, 2021 (Control 2021).

### Oceania

Oceania has total COVID-19 positive cases till May 15^th^, 2021 are 33454.0 and recover cases are 31901.0. The death ratio of the COVID-19 in Oceania was 2.8636%. Figure-4 (F) shows the recover cases and positive cases. The total test occur was 11088450.0 so the ration of positive cases among tests are 0.3017%. Figure-5 (F) is showing the graph cases occur from 26th January 2020 to May 15^th^, 2021.

### Effect of COVID-19 Vaccine and out of vaccination campaigns

The COVID-19 Vaccine Program in the United States was started on December 14, 2020. About 87.49 million individuals, or 26.4% of the population of the United States, have received at least one dose of vaccine as of March 26, 2021. Around 39.9 million people, or 12.07% of the US population, have received fully vaccination. Figure-6 (A) is showing that still 73.6 % US population requires COVID-19 vaccination. Figure-6 (B) and (C) (line chart) reveals that after Jan-2021 the chart of new cases is curve and new cases are decreasing day by day. Mostly there are three kind of vaccines available in the USA namely Pfizer-BioNTech, Moderna And J&J (Control 2021).

**Figure-6.**
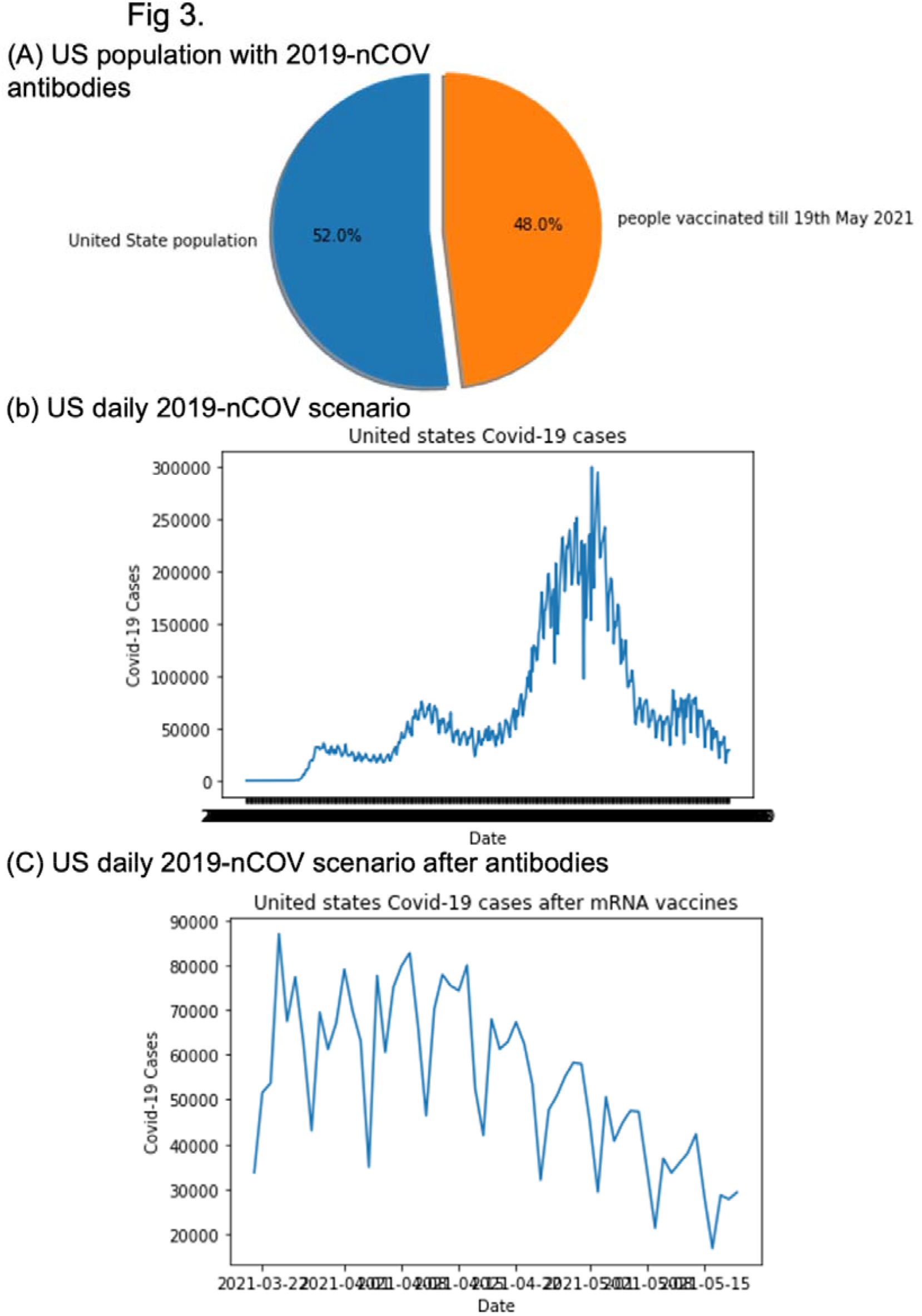
A comparison of case-based antibodies of COVID-19 observed in the U.S.

## Conclusion

The current work represents a novel code - scripting big data analysis of COVID-19 using Apache Spark and Python, as well as a machine learning approach that any researcher can use.

The advantage of this study is real time data set rows were examined and endeavoured at the speed of microseconds of time. These data comprised of six distinct continents of the world. Based on this data we concluded the death ratio, positive COVID cases rate changes were observed very thoroughly, Although Africa has the lowest death rate, it is not statistically significantly lower than other continents. Interestingly, we observed that the Oceanian continent has a higher death rate than the other continents, even though the total number of COVID testing is very low. In Asia, 6% of the total COVID testing population tested positive for COVID, whereas 9% of the total COVID testing population tested positive for COVID in North America. Even though the total number of COVID testing population in North as well South America is greater than either Asia or other continents. We also observed that after vaccination, positive COVID cases begin to decrease dramatically, which is a result of the ongoing successful vaccination campaign program in the United States and the North American continent (Figure-6). In Conclusion, this Apache Spark and Python based a machine learning approach implements the time dependent data availability with live coverage of updates with personal hygiene and precautions towards COVID-19. Represented machine learning-based PySpark analysis is also useful for healthcare research institutes in clinical and biostatistics rapid analysis, allowing them to conduct a quick, efficient, visualizing, and agile study on the COVID-19 outbreak.

## Data Availability

https://github.com/owid/covid-19-data/blob/master/public/data

## Compliance with Ethical Standards

The authors declare that they have no known competing financial interests or personal relationships that could have appeared to influence the work reported in this paper.

This article does not contain any studies with human participants or animals performed by any of the authors.

This work relies on the online data available from Center for Systems Science and Engineering (CSSE) at Johns Hopkins University.

